# History of suicide attempts and COVID-19 infection in Veterans with schizophrenia or schizoaffective disorder: effect modification by age and obesity

**DOI:** 10.1101/2021.08.25.21262627

**Authors:** Olaoluwa O. Okusaga, Rachel L. Kember, Gina M. Peloso, Roseann E. Peterson, Mariana Vujkovic, Brian G. Mitchell, Jared Bernard, Annette Walder, Tim B. Bigdeli

**Affiliations:** Mental Health Care Line, Michael E. DeBakey VA Medical Center, Houston, TX, USA; Menninger Department of Psychiatry, Baylor College of Medicine, Houston, TX, USA; Department of Psychiatry, Perelman School of Medicine, University of Pennsylvania, Philadelphia, PA, USA; Corporal Michael J. Crescenz VA Medical Center, Philadelphia, PA, USA; Department of Biostatistics, Boston University School of Public Health, Boston MA, USA; Virginia Institute for Psychiatric and Behavioral Genetics, Department of Psychiatry, Virginia Commonwealth University, Richmond, VA, USA; Department of Medicine, University of Pennsylvania Perelman School of Medicine, Philadelphia, PA, USA; Center for Innovations in Quality, Effectiveness and Safety (IQuEST), Michael E. DeBakey VA Medical Center, Houston, TX; VA New York Harbor Healthcare System, Brooklyn, NY; Department of Psychiatry and Behavioral Sciences, SUNY Downstate Medical Center, Brooklyn, NY

**Author notes:** Corresponding Author: Olaoluwa O. Okusaga, MD, MScPHR, Michael E. DeBakey Veterans Affairs Medical Center, 2002 Holcombe Boulevard, 580/116 MHCL rTMS, Houston, Texas, 77030.

## Abstract

**Importance:** As patients with schizophrenia or schizoaffective disorder have a high risk of suicide, and a history of suicide attempt is a strong predictor of suicide, determining whether history of suicide attempt is associated with COVID-19 in patients with schizophrenia or schizoaffective disorder has implications for suicide prevention in this patient population.

**Objective:** To determine whether a history of suicide attempt is associated with COVID-19 in Veterans with schizophrenia or schizoaffective disorder.

**Design:** Cross-sectional analyses of nation-wide electronic health records (EHR).

**Setting:** United States Veterans Health Administration.

**Participants:** Veterans with a diagnosis of schizophrenia or schizoaffective disorder that received treatment at any United States Veterans Affairs Medical Center from January 1, 2020 to January 31, 2021.

**Exposure:** History of suicide attempt.

**Main Outcome:** Adjusted and unadjusted odds ratios (ORs) for COVID-19 positivity in suicide attempters relative to non-attempters. Adjusted analyses included age, sex, race, marital status, BMI, and a medical comorbidity score.

**Results:** A total of 101,032 Veterans [mean age 56.67 ± 13.13 years; males 91,715 (90.8%)] were included in the analyses. There were 2,703 (2.7%) suicide attempters and 719 (0.7%) patients were positive for COVID-19. There was effect modification by age and BMI in the association of history of suicide attempt with COVID-19 positivity such that the association was only significant in obese (BMI ≥ 30) patients and patients younger than 59 years respectively. In the entire sample, the unadjusted OR for COVID-19 positivity in attempters was 1.42 (95% CI 0.97 to 2.10) and the adjusted odds ratio was 1.90 (95% CI 1.28 to 2.80). In patients younger than 59 years, and in the obese patients respectively, history of suicide attempt was associated with COVID-positive status in unadjusted analyses [OR 3.53 (95% CI 2.10 to 5.94); OR 2.22 (95% CI 1.29 to 3.81)] and adjusted analyses [OR 3.42 (95% CI 2.02 to 5.79); OR 2.85 (95% CI 1.65 to 4.94)].

**Conclusions and Relevance:** Young or obese suicide attempters with a diagnosis of schizophrenia or schizoaffective disorders have higher rates of COVID-19 diagnosis; due to possible long-term neuropsychiatric sequelae of infection with SARS-CoV-2, such patients should be monitored closely.

---

The risk of suicide in patients with schizophrenia is estimated to be 4.5 times higher than that for the general US population.^1^ A history of military service is also associated with increased risk of suicide, and the rate of suicide in US Veterans was 1.5 times that of the general US adult population from 2013 to 2018.^2^ Importantly, a diagnosis of schizophrenia further increases the risk of suicide in Veterans, and suicide mortality rate as high as 16.3% have been reported in male Veterans with schizophrenia.^3^ Although clinicians have a poor ability to predict suicide, a history of suicide attempt is significantly associated with future suicide.^4^ Therefore, characterizing biological outcomes in patients with a history of suicide attempt could lead to the identification of biomarkers^5^ (indicators of pathogenic processes) of suicidal behavior and further our understanding of suicide.

One biological factor with potential for biomarker function in patients with a history of suicide attempt, is infection. Increased risk of death from suicide, in a dose-response relationship, has previously been reported in individuals hospitalized for infection,^6^ and a parasitic infection has been associated with suicide attempts in patients with schizophrenia,^7^ but no study has evaluated the association of history of suicide attempt with infection in patients with schizophrenia or schizoaffective disorder. Moreover, immune activation and inflammation (which can be triggered by infection) have been found to be dysregulated in suicide attempters relative to non-attempters.^8,9^ Consequently, we sought to determine whether a history of suicide attempt in US Veterans diagnosed with schizophrenia or schizoaffective disorder is associated with testing positive for COVID-19. COVID-19 is caused by the novel severe acute respiratory syndrome coronavirus 2 (SARS-CoV-2) and has thus far claimed over 3 million lives globally since it was first described in December 2019.^10^ Since dimensions and behavioral traits (e.g. anhedonia, cognitive deficits and impulsivity) associated with suicidal behavior^11^ have also been associated with reduced propensity to adopt COVID-19 precautionary behaviors,^12^ we hypothesized that a history of suicide attempt would be associated with COVID-19 positivity in Veterans with schizophrenia or schizoaffective disorder.

## Methods

### Study Design, Setting and Participant

A cross-sectional design was employed, involving analyses of nation-wide electronic health records (EHR) of a large sample of Veterans with a diagnosis of schizophrenia or schizoaffective disorder that received treatment at any United States Veterans Affairs Medical Center (VAMC) from January 1, 2020 to January 31, 2021. Approval for the study was granted by the Institutional Review Board (IRB) of Baylor College of Medicine and the Michael E. DeBakey VAMC Research and Development Committee. The IRB granted a waiver of consent and HIPAA authorization since the use of deidentified protected health information involved no more than minimal risk (including privacy risks) to the individuals in this study.

### Data Collection and Definitions

Data were collected from the Corporate Data Warehouse (CDW) and the Veterans Affairs Informatics and Computing Interface (VINCI).^13^ CDW incorporates data from several data sets (including EHR) throughout the Veteran’s Health Administration into one standard database structure to facilitate reporting and data analysis. VINCI is a partner with the CDW and hosts all data available through CDW. However, if a Veteran receives care at a non-VA healthcare facility, then the data for that encounter will not automatically be captured by CDW/VINCI.

The International Classification of Diseases, Ninth and Tenth Revisions codes (ICD-9 and ICD-10 codes) were used to identify Veterans with a diagnosis of schizophrenia or schizoaffective disorder. ICD-9 and ICD-10 codes were also used to identify patients with a documented suicide attempt. The ICD-9 and ICD-10 codes are provided in eTable 1 of the supplementary material. The outcome of interest, COVID-19-positive test results, were also retrieved from the EHR. Data regarding the specific type of COVID-19 test was not obtained. Veterans can request COVID-19 testing at no personal cost, but COVID-19 tests are not routinely ordered if a Veteran is not admitted to an inpatient unit or undergoing diagnostic or therapeutic procedures such as electroconvulsive therapy (ECT). The following variables were also extracted for each research subject: age (at last visit), sex, race, marital status, Body Mass Index (BMI), and Charlson-Deyo Score.^14^ The Charlson-Deyo Comorbidity Index is a clinical comorbidity index which assigns a score to the various chronic medical conditions present in an individual patient and the sum of the scores is predictive of long-term mortality.^15^

**Table 1.** Characteristics of the total sample of Veterans with schizophrenia or schizoaffective disorder, suicide attempters, and non-attempters.

| Characteristics | Total sample<br>(n=101,032)) | Attempters<br>(n=2703) | Non-attempters<br>(98,329) |
| --- | --- | --- | --- |
| Age (SD) | 56.67 ± 13.13 | 53.43 ± 12.58 | 56.76 ± 13.13 |
| Sex (n, %) |  |  |  |
| Male | 91,715 (90.8)) | 2349 (86.9) | 89,366 (90.9) |
| Female | 9,317 (9.2) | 354 (13.1) | 8,963 (9.1) |
| Race (n, %) |  |  |  |
| White | 55,941 (55.4) | 1,765 (65.3) | 54,176 (55.1) |
| Black | 35,589 (35.2) | 756 (28.0) | 34,833 (35.4) |
| Asian | 1,114 (1.1) | 22 (0.8) | 1,092 (1.1) |
| American Indian<br>or Alaska native | 1,003 (1.0) | 34 (1.3) | 969 (1.0) |
| Native Hawaiian<br>or other Pacific<br>Islander | 1155 (1.1) | 23 (0.9) | 1,132 (1.2) |
| Unknown | 6,230 (6.2) | 103 (3.8) | 6,127 (6.2) |
| Marital Status (n, %) |  |  |  |
| Married | 22,983 (22.7) | 480 (17.8) | 22,503 (22.9) |
| Divorced | 29,656 (29.4) | 1,029 (38.1) | 28,627 (29.1) |
| Never married | 38,671 (38.3) | 918 (34.0) | 37,753 (38.4) |
| Separated | 5,604 (5.5) | 165 (6.1) | 5,439 (5.5) |
| Widowed | 3,439 (3.4) | 103 (3.8) | 3,336 (3.4) |
| Single | 223 (0.2) | 6 (0.2) | 217 (0.2) |
| Unknown | 456 (0.5) | 2 (0.1) | 454 (0.5) |
| BMI Category (n, %) |  |  |  |
| Obese | 40,717 (40.3) | 1,069 (39.5) | 39,648 (40.3) |
| Not obese | 60,315 (59.7) | 1,634 (60.5) | 58,681 (59.7) |
| Total Deyo Score (SD) | 0.68 ± 1.42 | 0.82 ± 1.57 | 0.67 ± 1.41 |

BMI=body mass index. Total Deyo Score is an indicator of long-term mortality and higher scores indicate higher medical comorbidities.

### Statistical Analysis

This sample of Veterans with schizophrenia or schizoaffective disorder were divided into two groups—suicide attempters and non-attempters. Demographic and clinical variables were compared between attempters and non-attempters using t-tests or chi-squared as appropriate. Means ± standard deviation (SD) and n (%) are reported. Logistic regression models were applied to estimate the odds of testing positive for COVID-19 and comparing attempters to non-attempters, with additional analysis adjusting for age, sex, race, marital status, BMI and total Charlson-Deyo score. Based on previous findings of more severe COVID-19 illness in the elderly^16^, obese^17^ and those with comorbid medical conditions^18^, we also fitted COVID status-by-age, COVID status-by-BMI, and COVID status-by-Charlson-Deyo-score interactions in logistic regression models. COVID status-by-age, and COVID status-by-BMI interactions were significant (B= -0.018, p<0.001 and B= 0.772, p=0.05 respectively) but COVID status-by-Charlson-Deyo score was not significant (B= -0.042, p=0.612). We therefore carried out subgroup analyses in patients younger than 59 years (the median age of the sample), patients older than 59 years, obese (BMI ≥ 30), and non-obese patients (BMI<30) respectively. P-values less than 0.05 were considered statistically significant. All statistical analyses were performed with IBM SPSS, Version 27 (IBM Corp., Armonk, NY, USA).

## Results

A total of 101,032 Veterans [mean age 56.67 ± 13.13 years; males 91,715 (90.8%)] with either schizophrenia or schizoaffective disorder were included in the analyses. 2,703 (2.7%) patients had a history of suicide attempt. Table 1 shows the demographic and clinical characteristics of suicide attempters and non-attempters. Mean age of suicide attempters was 53.43 ± 12.58 years and mean age of non-attempters was 56.76 ± 13.13. 86.9% of the suicide attempters were male while 90.9% of the non-attempters were male. There was a total of 719 (0.7%) COVID-positive cases in the entire sample. Table 2 shows the demographic and clinical characteristics of the COVID-positive and Not COVID-positive groups respectively.

**Table 2.**
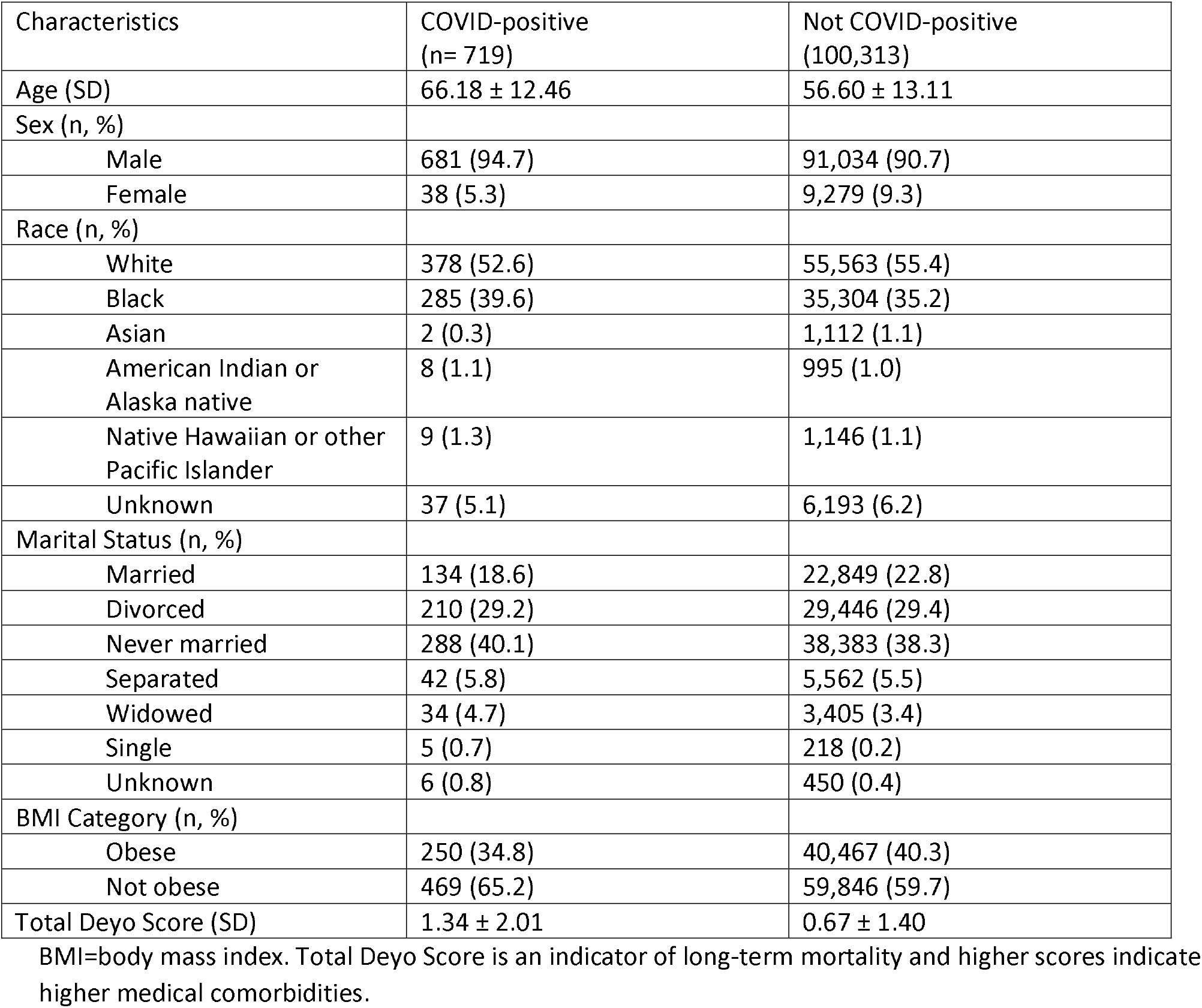
Characteristics of COVID-positive and Not COVID-positive Veterans with schizophrenia or schizoaffective disorder.

### History of suicide attempt and COVID-19 positivity in the entire sample

There was a higher percentage of COVID-positive cases in attempters relative to non-attempters (Table 3), but this was not statistically significant (1.0 % vs. 0.7%, χ^2^ = 3.24, p= 0.072). The unadjusted odds ratio for COVID positivity in patients with a history of suicide attempt was 1.42 (95% CI 0.97 to 2.10) and the adjusted odds ratio was 1.90 (95% CI 1.28 to 2.80).

**Table 3.** Number and percentage of COVID-positive cases based on suicide attempter/non-attempter status in the total sample and the subgroups of Veterans with schizophrenia or schizoaffective disorder.

| **Group** | **COVID-positive cases (n, %) in attempters** | **COVID-positive cases (n, %) in non-attempters** | **P-value\*** |
| --- | --- | --- | --- |
| Total sample (n= 101,032) | 27 (1.0) | 692 (0.7) | 0.072 |
| Younger than 59 years (n=50,490) | 16 (1.0) | 137 (0.3) | <0.001 |
| Older than 59 years (n=50,542) | 11 (1.0) | 555 (1.1) | 0.884 |
| Obese (40,717) | 14 (1.3) | 236 (0.6) | 0.008 |
| Not obese (60,315) | 13 (0.8) | 456 (0.8) | 0.886 |

\*Fisher's Exact Test

### Results of subgroup analyses

In patients younger than 59 years, and in the obese patients respectively, history of suicide attempt was associated with COVID-positive status in unadjusted analyses [OR 3.53 (95% CI 2.10 to 5.94); OR 2.22 (95% CI 1.29 to 3.81)] and adjusted analyses [OR 3.42 (95% CI 2.02 to 5.79); OR 2.85 (95% CI 1.65 to 4.94)]. In patients older than 59 years, and in the non-obese patients, there was no statistically significant association between history of suicide attempt and COVID-19 positivity from the unadjusted and adjusted analyses (table 4).

**Table 4.** Odds ratios for COVID-19 positivity based on suicide attempt status (attempters vs non-attempters) in the total sample and subgroup of Veterans with schizophrenia or schizoaffective disorder

| **Group** | **Unadjusted Odds Ratio with 95% CI** | **Adjusted Odds Ratio with 95% CI\*** |
| --- | --- | --- |
| Total Sample (n = 101,032) | 1.42 (0.97-2.10) | 1.90 (1.28-2.80) |
| Younger than 59 years (n = 50,490) | 3.53 (2.10-5.94) | 3.42 (2.02-5.79) |
| Older than 59 years (n = 50,542) | 0.91 (0.50-1.66) | 0.91 (0.50-1.66) |
| Obese (n = 40,717) | 2.21 (1.29-3.81) | 2.85 (1.65-4.94) |
| Not-obese (n = 60,315) | 1.02 (0.59-1.78) | 1.39 (0.80-2.43) |

\*Adjusted for age, sex, race, marital status, BMI and total Charlson-Deyo score.

## Discussion

In this nationwide Veteran’s Health Administration study, a history of suicide attempt was associated with testing positive for COVID-19 in Veterans with schizophrenia or schizoaffective disorder who were either younger than 59 years or obese. In the entire sample, the unadjusted odds ratio for the association of history of suicide attempt with COVID positivity was not statistically significant but the adjusted odds ratio was significant, a finding likely due to negative confounding (i.e. significant association of a covariate with the outcome).^19^ To our knowledge, this is the first study to report an association between history of suicide attempt and COVID-19 in Veterans with schizophrenia or schizoaffective disorder.

The association of history of suicide attempt with COVID-19 positivity in the younger and obese subgroups is noteworthy considering previous research findings. For example, suicide risk in schizophrenia is highest for young adults but lowest for patients older than 65 years^1^ and although BMI has not been associated with suicide risk in schizophrenia or schizoaffective disorder, obesity is associated with higher levels of immune system dysregulation and inflammation in patients with schizophrenia.^20,21^ Relatedly, markers of immune dysregulation and inflammation in the periphery and central nervous system are elevated in suicide attempters relative to non-attempters.^8,9^ Furthermore, primary humoral immunodeficiencies were recently robustly associated with psychiatric disorders (including schizophrenia) and suicide attempts in the same cohort.^22^ It is, therefore, conceivable that the association of suicide attempt history with COVID-19 might reflect dysregulated immune function in young and obese attempters respectively, since individuals with conditions involving immune system dysregulation such as asthma and allergic rhinitis, are more likely to test positive for COVID-19^23^ relative to those without such conditions.

Another possible explanation of the association of history of suicide attempt with COVID-19 in Veterans with schizophrenia or schizoaffective disorder is that psychological factors which predispose patients to suicide attempts also make them more vulnerable to COVID-19 infection. For example, anhedonia, impulsivity and substance abuse are associated with suicidal behavior and have also been associated with less likelihood of adopting COVID-19 precautionary behaviors such as physical distancing, wearing mask or hand-washing.^12,24^

Due to possible long-term neuropsychiatric sequelae of infection with SARS-CoV-2,^25^ patients with schizophrenia or schizoaffective disorder who have a history of suicide attempt and are younger than 59 or obese, should be monitored closely since they are more at risk of COVID-19. Indeed, there is the possibility of symptom exacerbation in patients with a history of suicide attempt who test positive for COVID-19 since post-mortem examination of brains of individuals diagnosed with severe COVID-19 infection revealed COVID-19-related impaired brain neurotransmission.^26^ Furthermore, COVID-19 differentially expressed genes in the patients’ brains were enriched in genetic variants associated with schizophrenia and a schizophrenia-related trait (cognition) in a cell type-specific manner.^26^.

This study is subject to several limitations. First, the cross-sectional design makes it impossible to infer a causal association between suicide attempt and COVID-19. Second, we do not have data on the specific nature and severity of suicide attempts. Third, the sample was predominantly male which limits the generalizability of our results. Fourth, we were unable to adjust for psychotropic medications, especially clozapine which has been associated with increased likelihood of testing positive for COVID-19.^27^ Finally, we did not have information on illness severity and so did not adjust for it. Related to the last limitation is our inability to control for inpatient vs. outpatient status since inpatients are more likely to get tested for COVID.

In summary, our findings provide preliminary evidence of an association between history of suicide attempt and COVID-19 in a subgroup of patients with schizophrenia or schizoaffective disorder and therefore support the expansion of strategies to ensure vaccination of patients with schizophrenia or schizoaffective disorder, especially given the fact that patients with schizophrenia are under vaccinated against COVID-19.^28^ Future well-powered, prospective studies are needed to confirm our observed association between history of suicidal behavior and COVID-19 in patients with schizophrenia or schizoaffective disorder and also to elucidate the underlying mechanisms of the association.

## Data Availability

The data that support the findings of this study are not publicly available, but a minimal data set required to replicate the study findings reported in the article will be made available on request to the corresponding author.

---

**eTable 1:** ICD codes for schizophrenia/schizoaffective disorder, suicide attempt and COVID-19.

| Diagnosis | ICD-9 | ICD-10 |
| --- | --- | --- |
| Schizophrenia and schizoaffective disorder | 295, 295.0, 295.1, 295.2, 295.3<br>295.4, 295.5, 295.6, 295.7,<br>295.8, 295.9 | F20, F20.0, F20.1, F20.2, F20.3<br>F20.5, F20.8, F20.89, F20.9, F25,<br>F25.0, F25.1, F25.8, F25.9 |
| Suicide attempt | E950, E951, E952, E953, E954,<br>E955, E956, E957, E958, E959 | X71, X72, X73, X74, X75, X76,<br>X77, X78, X79, X80, X81, X82<br>X83, T36-T50, T51-T65, S51,<br>S55, S59, S61, S65, S69 |
| Confirmed diagnosis of the 2019 novel coronavirus (COVID19) |  | U07.1, COVID-19 |

## References

1. Olfson M, Stroup TS, Huang C, Wall MM, Crystal S, Gerhard T. Suicide Risk in Medicare Patients With Schizophrenia Across the Life Span. JAMA Psychiatry. Published online May 26, 2021. doi:10.1001/jamapsychiatry.2021.0841

2. Sokol Y, Gromatsky M, Edwards ER, et al. The deadly gap: Understanding suicide among veterans transitioning out of the military. Psychiatry Res. 2021;300. doi:10.1016/j.psychres.2021.113875

3. Aslan M, Radhakrishnan K, Rajeevan N, et al. Suicidal ideation, behavior, and mortality in male and female US veterans with severe mental illness. J Affect Disord. 2020;267:144–152. doi:10.1016/j.jad.2020.02.022

4. Bostwick JM, Pabbati C, Geske JR, McKean AJ. Suicide attempt as a risk factor for completed suicide: Even more lethal than we knew. Am J Psychiatry. 2016;173(11):1094–1100. doi:10.1176/appi.ajp.2016.15070854

5. Atkinson AJ, Colburn WA, DeGruttola VG, et al. Biomarkers and surrogate endpoints: Preferred definitions and conceptual framework. Clin Pharmacol Ther. 2001;69(3):89–95. doi:10.1067/mcp.2001.113989

6. Lund-Sørensen H, Benros ME, Madsen T, et al. A Nationwide Cohort Study of the Association Between Hospitalization With Infection and Risk of Death by Suicide. JAMA Psychiatry. 2016;70(8):812–820. doi:10.1001/jamapsychiatry.2016.1594

7. Okusaga O, Langenberg P, Sleemi A, et al. Toxoplasma gondii antibody titers and history of suicide attempts in patients with schizophrenia. Schizophr Res. 2011;133(1-3):150–155. doi:10.1016/j.schres.2011.08.006

8. Ducasse D, Olié E, Guillaume S, Artéro S, Courtet P. A meta-analysis of cytokines in suicidal behavior. Brain Behav Immun. 2015;46:203–211. doi:10.1016/j.bbi.2015.02.004

9. Ganança L, Oquendo MA, Tyrka AR, Cisneros-Trujillo S, Mann JJ, Sublette ME. The role of cytokines in the pathophysiology of suicidal behavior. Psychoneuroendocrinology. 2016;63:296–310. doi:10.1016/j.psyneuen.2015.10.008

10. Osuchowski MF, Winkler MS, Skirecki T, et al. The COVID-19 puzzle: deciphering pathophysiology and phenotypes of a new disease entity. Lancet Respir Med. 2021;9(6):622–642. doi:10.1016/s2213-2600(21)00218-6

11. Turecki G, Brent DA, Gunnell D, et al. Suicide and suicide risk. Nat Rev Dis Prim. 2019;5(1). doi:10.1038/s41572-019-0121-0

12. Frías-Armenta M, Corral-Frías NS, Corral-Verdugo V, Lucas MY. Psychological Predictors of Precautionary Behaviors in Response to COVID-19: A Structural Model. Front Psychol. 2021;12. doi:10.3389/fpsyg.2021.559289

13. Price LE, Shea K, Gephart S. The Veterans Affairs’s Corporate Data Warehouse: Uses and Implications for Nursing Research and Practice. Nurs Adm Q. 2015;39(4):311–318. doi:10.1097/NAQ.0000000000000118

14. Deyo RA, Cherkin DC, Ciol MA. Adapting a clinical comorbidity index for use with ICD-9-CM administrative databases. J Clin Epidemiol. 1992;45(6):613–619. doi:10.1016/0895-4356(92)90133-8

15. Ladha KS, Zhao K, Quraishi SA, et al. The Deyo-Charlson and Elixhauser-van Walraven Comorbidity Indices as predictors of mortality in critically ill patients. BMJ Open. 2015;5(9). doi:10.1136/bmjopen-2015-008990

16. Mueller AL, Mcnamara MS, Sinclair DA. Why does COVID-19 disproportionately affect older people? Aging (Albany NY). 2020;12(10):9959–9981. doi:10.18632/aging.103344

17. Gonçalves DA, Ribeiro V, Gualberto A, Peres F, Luconi M, Gameiro J. COVID-19 and Obesity: An Epidemiologic Analysis of the Brazilian Data. Int J Endocrinol. 2021;2021:1–10. doi:10.1155/2021/6667135

18. Rosenthal N, Cao Z, Gundrum J, Sianis J, Safo S. Risk Factors Associated with In-Hospital Mortality in a US National Sample of Patients with COVID-19. JAMA Netw Open. 2020;3(12). doi:10.1001/jamanetworkopen.2020.29058

19. Mehio-Sibai A, Feinleib M, Sibai TA, Armenian HK. A positive or a negative confounding variable? A simple teaching aid for clinicians and students. Ann Epidemiol. 2005;15(6):421–423. doi:10.1016/j.annepidem.2004.10.004

20. Boozalis T, Devaraj S, Okusaga OO. Correlations between Body Mass Index, Plasma High-Sensitivity C-Reactive Protein and Lipids in Patients with Schizophrenia. Psychiatr Q. 2019;90(1):101–110. doi:10.1007/s11126-018-9606-3

21. Sirota P, Hadi E, Djaldetti M, Bessler H. Difference in inflammatory cytokine production by mononuclear cells from obese and non-obese schizophrenic patients. Acta Psychiatr Scand. 2015;132(4):301–305. doi:10.1111/acps.12396

22. Isung J, Williams K, Isomura K, et al. Association of Primary Humoral Immunodeficiencies with Psychiatric Disorders and Suicidal Behavior and the Role of Autoimmune Diseases. JAMA Psychiatry. 2020;77(11):1147–1154. doi:10.1001/jamapsychiatry.2020.1260

23. Yang JM, Koh HY, Moon SY, et al. Allergic disorders and susceptibility to and severity of COVID-19: A nationwide cohort study. J Allergy Clin Immunol. 2020;146(4):790–798. doi:10.1016/j.jaci.2020.08.008

24. Reinders Folmer C, Kuiper ME, Olthuis E, et al. Sustaining Compliance with COVID-19 Mitigation Measures? Understanding Distancing Behavior in the Netherlands during June 2020. SSRN Electron J. Published online September 2, 2020. doi:10.2139/ssrn.3682479

25. Kappelmann N, Dantzer R, Khandaker GM. Interleukin-6 as potential mediator of long-term neuropsychiatric symptoms of COVID-19. Psychoneuroendocrinology. 2021;131:105295. doi:10.1016/j.psyneuen.2021.105295

26. Yang AC, Kern F, Losada PM, et al. Dysregulation of brain and choroid plexus cell types in severe COVID-19. Nature. Published online June 21, 2021. doi:10.1038/s41586-021-03710-0

27. Govind R, Fonseca de Freitas D, Pritchard M, Hayes RD, MacCabe JH. Clozapine treatment and risk of COVID-19 infection: retrospective cohort study. Br J Psychiatry. Published online July 27, 2020:1-7. doi:10.1192/bjp.2020.151

28. Bitan DT. Patients with schizophrenia are under-vaccinated for COVID-19: a report from Israel. World Psychiatry. 2021;20(2):300–301. doi:10.1002/wps.20874

